# mRNA booster immunization elicits potent neutralizing serum activity against the SARS-CoV-2 Omicron variant

**DOI:** 10.1101/2021.12.14.21267769

**Authors:** Henning Gruell, Kanika Vanshylla, Pinkus Tober-Lau, David Hillus, Philipp Schommers, Clara Lehmann, Florian Kurth, Leif E. Sander, Florian Klein

## Abstract

The Omicron variant of SARS-CoV-2 is causing a rapid increase in infections in various countries. This new variant of concern carries an unusually high number of mutations in key epitopes of neutralizing antibodies on the spike glycoprotein, suggesting potential immune evasion. Here we assessed serum neutralizing capacity in longitudinal cohorts of vaccinated and convalescent individuals, as well as monoclonal antibody activity against Omicron using pseudovirus neutralization assays. We report a near-complete lack of neutralizing activity against Omicron in polyclonal sera after two doses of the BNT162b2 vaccine, in convalescent individuals, as well as resistance to different monoclonal antibodies in clinical use. However, mRNA booster immunizations in vaccinated and convalescent individuals resulted in a significant increase of serum neutralizing activity against Omicron. Our study demonstrates that booster immunizations will be critical to substantially improve the humoral immune response against the Omicron variant.

## Main

Most approved COVID-19 vaccines are based on transient expression of the viral spike (S) glycoprotein (Wu01 strain) to induce SARS-CoV-2-directed immunity.^1^ Mutations in antibody epitopes on the spike protein can result in increased viral resistance to neutralizing antibodies and have been associated with reduced vaccine effectiveness.^2^ Moreover, they are able to strongly impair activity of monoclonal antibodies used for treatment and prevention of COVID-19.^3^

Shortly after its identification in the Gauteng province in South Africa, the Omicron variant (BA.1 sublineage of B.1.1.529) of SARS-CoV-2 was designated as a “variant of concern” (VoC) by the World Health Organization. Genomic surveillance and surrogate parameters (e.g., S gene target failure in diagnostic PCR) document a sharp rise in Omicron infections across the globe.^4^ Moreover, the observation of increasing incidences in populations with high prevalence of SARS-CoV-2 immunity as well as reports of re-infections suggest potent immune evasion properties of the Omicron variant.^5^ In comparison to previously described VoCs, the Omicron variant is notable for its high number of non-synonymous mutations relative to the ancestral Wu01 strain of SARS-CoV-2. The majority of these mutations are located in the viral spike glycoprotein and include critical epitopes for SARS-CoV-2-neutralizing antibodies in the N-terminal domain and the receptor-binding domain. Several of these mutations have already been associated with resistance to SARS-CoV-2- or vaccine-induced neutralizing antibodies.^3,6,7^ Therefore, the spread of the Omicron variant can have important implications on current strategies to prevent and treat COVID-19, and may require urgent public health interventions to limit transmission and morbidity.

To determine the susceptibility of the Omicron variant to vaccine-induced serum activity, we analyzed samples obtained from 30 individuals with no evidence of prior infection.^8^ Samples were collected one month (median 3.9 weeks; range 2.9-6.0 weeks) after completion of a two-dose course of the BNT162b2 vaccine (**Fig. 1a**). Study participants had a median age of 49 years (range 27-78 years) with nearly equal sex distribution (57% females, 43% males; **Supplementary Table 1**). Sera were tested in a lentivirus-based neutralization assay using pseudoviruses expressing the spike proteins of the Wu01 vaccine strain, or the Alpha (B.1.1.7), Beta (B.1.351), Delta (B.1.617.2), or Omicron VoC (**Fig. 1a** and **Supplementary Table 2**). All samples showed high levels of neutralizing activity against the Wu01 strain with a geometric mean 50% inhibitory serum dilution (GeoMean ID_50_) of 546 (**Fig. 1a**). Serum neutralizing activity against the Alpha, Delta, and Beta variant was decreased to GeoMean ID_50_s of 331, 172, and 40, respectively (samples that did not achieve 50% inhibition at the lowest tested dilution of 10 were imputed to an ID_50_ of 5). Notably, only eight out of the 30 vaccinated individuals (27%) displayed detectable serum neutralizing activity against Omicron, resulting in a GeoMean ID_50_ of 8 (**Fig. 1a**), which was significantly lower than against the Beta variant (*P*<0.0001), one of the most immune evasive variants previously described.^2^

**Figure 1.**
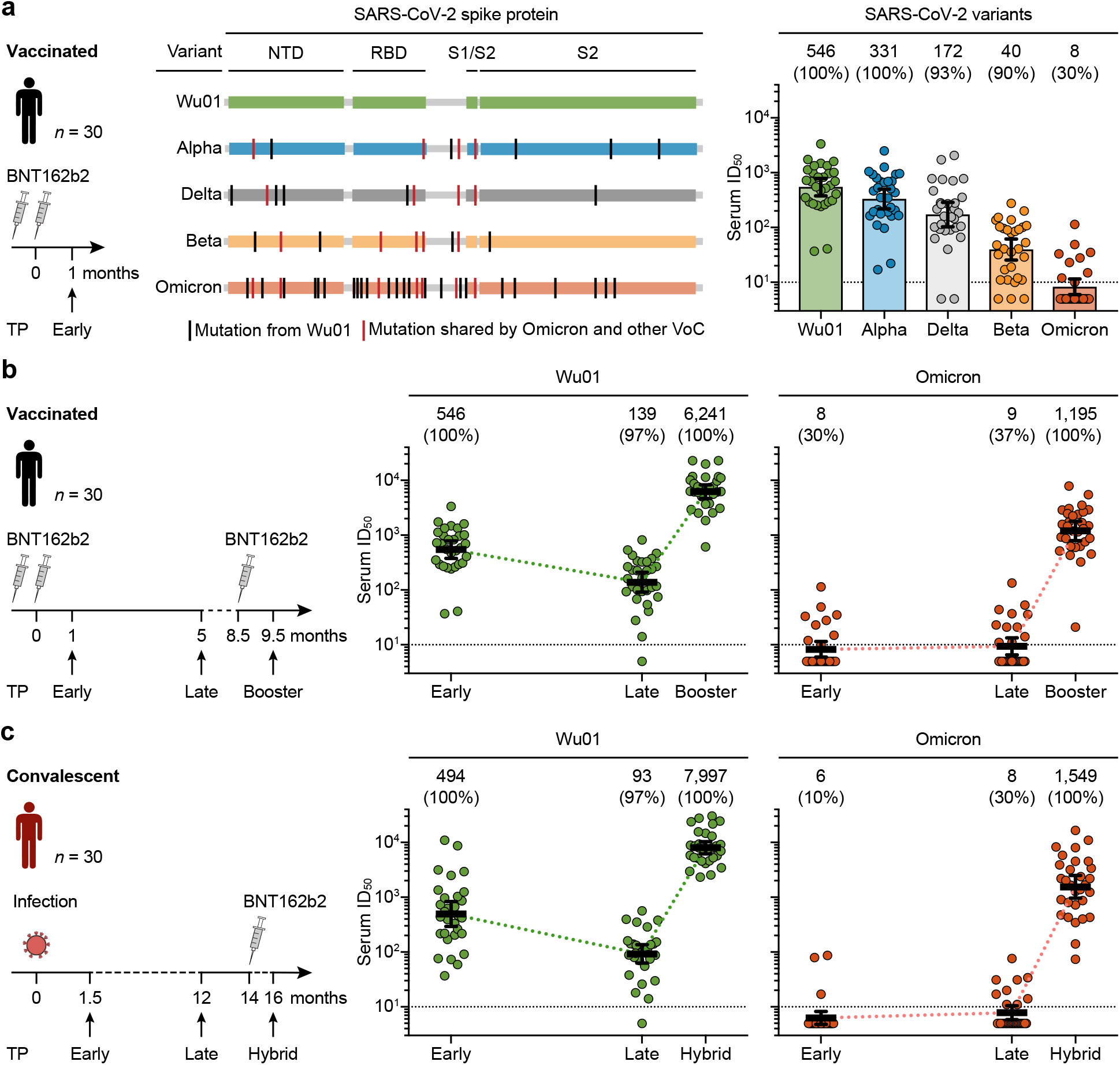
SARS-CoV-2-neutralizing serum activity in vaccinated and convalescent individuals. **a**, Neutralizing serum activity was determined in samples obtained one month after two doses of BNT162b2 against the ancestral Wu01 strain of SARS-CoV-2 and four variants of concern. Fifty-percent inhibitory serum dilutions (ID_50_s) were determined by pseudovirus neutralization assays. Bars indicate geometric mean ID_50_s with 95% confidence intervals (CIs). **b**, Serum ID_50_s against the Wu01 strain (left) and the Omicron variant (right) of SARS-CoV-2 in a longitudinal cohort of 30 individuals. Samples were collected at a median of one month (*Early*, see **a**) and five months (*Late*) after two doses of BNT162b2, and one month after a subsequent single dose of BNT162b2 (*Booster*). Colored lines connect geometric mean ID_50_s and error bars indicate 95% CIs. **c**, Serum ID_50_s against the Wu01 strain (left) and the Omicron variant (right) of SARS-CoV-2 in a longitudinal cohort of 30 COVID-19-convalescent individuals. Samples were collected at a median of 1.5 (*Early*) and 12 months (*Late*) after diagnosis of SARS-CoV-2 infection, as well as two months after a single BNT162b2 vaccination (*Hybrid*). Colored lines connect geometric mean ID_50_s and error bars indicate 95% CIs. In **a, b**, and **c**, ID_50_s below the lower limit of quantification (LLOQ, ID_50_ of 10; indicated by black dashed lines) were imputed to ½ LLOQ (ID_50_=5). Numbers above graphs indicate geometric mean ID50s and the percentage of samples with detectable neutralizing activity in parentheses. TP, time point; NTD, N-terminal domain; RBD, receptor-binding domain.

To investigate the change in serum neutralizing activity against the Omicron variant over time and assess the impact of a booster vaccination, we analyzed longitudinal samples of the 30 vaccinees. Neutralizing serum activity against Wu01 and Omicron was determined at a median of five months (*Late* time point) after BNT162b2 vaccination, as well as one month after a single BNT162b2 booster dose (*Booster* time point; **Fig. 1b**). Following two-dose BNT162b2, neutralizing activity against Wu01 decreased four-fold over the period of five months from a GeoMean ID_50_ of 546 to 139, but was strongly increased after booster vaccination (GeoMean ID_50_ of 6,241). Serum neutralizing activity against the Omicron variant remained low with only 30%-37% of the samples showing detectable neutralization, resulting in GeoMean ID_50_s of 8 and 9 at the *Early* and *Late* time points, respectively. However, neutralizing serum activity to the Omicron variant increased by more than 100-fold following the booster dose of BNT162b2 resulting in a GeoMean ID_50_ of 1,195, and was detectable in all 30 participants (100%) (**Fig. 1b**). Strikingly, serum neutralizing activity against the Omicron variant following booster immunization was even higher than neutralizing titers against Wu01 after two doses of BNT162b2 (GeoMean ID_50_ of 1,195 vs. 546; *P*=0.0003).

Moreover, we analyzed the neutralizing serum response to the Omicron variant in a longitudinal cohort of 30 COVID-19 convalescent individuals (**Supplementary Table 1**).^9^ Study participants were followed for up to 16 months from the time of SARS-CoV-2 infection which occurred between February and March 2020 (i.e., prior to the emergence of WHO-designated VoCs). Serum samples were collected at 1.5 months (range 3.6-10 weeks, *Early* time point) and 12 months (range 49.4-59.4 months, *Late* time point) after diagnosis of infection (**Fig. 1c**). In addition, serum was obtained one month after administration of a single dose BNT162b2 resulting in a “hybrid immunity” acquired by a combination of infection and vaccination (*Hybrid* time point; **Fig. 1c**). Early after infection, neutralizing activity against Wu01 was relatively variable, ranging in ID_50_s from 37 to 11,008, with a GeoMean ID_50_ of 494 that decreased to 93 after 12 months (*Late*). Following a single BNT162b2 dose, all convalescent individuals had a strong increase in neutralizing serum activity resulting in a GeoMean ID_50_ of 7,997 against Wu01 (*Hybrid*). No or only weak neutralizing activity was determined against the Omicron variant in samples collected from convalescent individuals at the *Early* and *Late* time points. However, a slight increase in neutralizing activity was observed at the *Late* time point in some individuals, potentially indicating ongoing affinity maturation resulting in antibodies capable of targeting a wider spectrum of SARS-CoV-2 variants (**Supplementary Table 2**). Despite the near-lack of neutralizing serum activity against the Omicron variant at a median of 12 months after SARS-CoV-2 infection (GeoMean ID_50_ of 8; *Late* time point), a single dose of BNT162b2 induced a remarkable increase demonstrated by a GeoMean ID_50_ of 1,549 one month after vaccination (*Hybrid* time point) (**Fig. 1c)**. We conclude that the Omicron variant exerts substantial humoral immune escape in BNT162b2-vaccinated and convalescent individuals. However, high levels of neutralizing activity against the Omicron variant can be induced by a BNT162b2 booster immunization.

Finally, we investigated whether the high number of mutations in the Omicron variant affects the activity of SARS-CoV-2-neutralizing monoclonal antibodies that have previously been demonstrated to effectively reduce COVID-19-associated morbidity and mortality.^10-12^ To this end, we determined the activity of nine monoclonal antibodies, including antibodies authorized for clinical use (bamlanvimab, etesevimab, REGN10933 (casirivimab), REGN10987 (imdevimab), and S309 (sotrovimab)) (**Table 1**). All antibodies were tested in parallel against the Wu01 strain as well as the Alpha, Beta, Delta, and Omicron variants. While all antibodies showed neutralizing activity against Wu01 and Alpha, only 7/9 and 5/9 showed neutralizing activity against the Delta and Beta variants, respectively (**Table 1**). Notably, neutralizing activity against the Omicron variant was abolished in seven out of nine antibodies (**Table 1**). We conclude that the neutralizing activity of several monoclonal antibodies is strongly affected against the Omicron variant and will limit treatment options for Omicron-induced COVID-19.

**Table 1.**
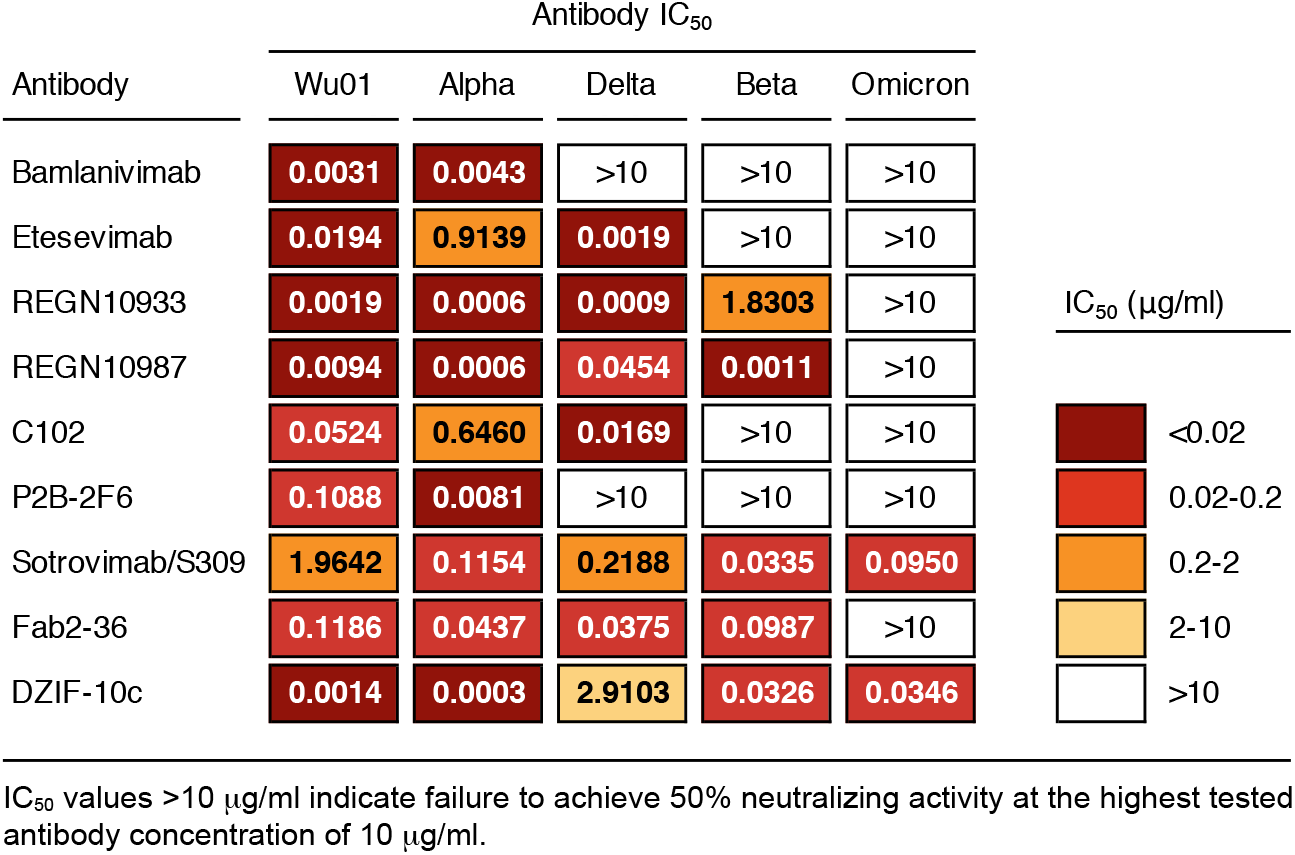
SARS-CoV-2-neutralizing activity of monoclonal antibodies.

The rapid surge in Omicron variant infections poses a significant challenge to public health.^4^ Our study demonstrates marked resistance of the Omicron variant to serum neutralizing activity induced by two doses of the BNT162b2 vaccine or SARS-CoV-2 infection. Lower neutralizing titers have been associated with an increased risk of symptomatic COVID-19, suggesting that limited neutralizing activity against Omicron may result in increased risk of infection and higher burden of disease.^13^ Whether cellular immunity will be effective in preventing severe disease after Omicron infection in the absence of a potent neutralizing antibody response remains to be determined.^14,15^ Importantly, a single BNT162b2 booster immunization effectively induced a substantial increase in serum neutralization against the Omicron variant and resulted in neutralizing titers similar to those observed against Wu01 after two doses of BNT162b2. It can be speculated that the extent of the potent neutralizing response against the Omicron variant following booster vaccination may be a consequence of ongoing affinity maturation following initial vaccination or SARS-CoV-2 infection with the ancestral spike protein.^16-18^ While variant-specific vaccines and novel monoclonal antibodies may be required for optimal activity, our data provide clear evidence that booster immunization with an available vaccine induces robust neutralization against the immune evasive Omicron variant. In addition to the critical need for rapidly making vaccines globally available and accessible to counter the emergence of novel variants, booster campaigns may provide an immediate and effective intervention against Omicron.

## Methods

### Study design

COVID-19-convalescent samples were obtained under protocols approved by the ethics committee (EC) of the Medical Faculty of the University of Cologne (16-054 and 20-1187). Individuals with PCR-confirmed SARS-CoV-2 infection (by anamnesis) were enrolled between April and May 2020 within eight weeks of diagnosis and/or symptom onset and followed longitudinally. Except for one hospitalized participant, all individuals reported having mild SARS-CoV-2-infection-related symptoms. Samples from non-infected vaccinated individuals were obtained under protocols (EICOV, COVIMMUNIZE, and COVIM) approved by the EC of Charité - Universitätsmedizin Berlin (EA4/245/20 and EA4/244/20; EICOV and COVIMMUNIZE), and by the Federal Institute for Vaccines and Biomedicines (Paul Ehrlich Institute) and the EC of the state of Berlin (COVIM). All samples were tested for the presence of anti-nucleocapsid antibodies using the SeraSpot Anti-SARS-CoV-2 IgG microarray-based immunoassay (Seramun Diagnostica). Individuals with a positive SARS-CoV-2 anamnesis or confirmed infection (positive nucleic acid amplification test or anti-nucleocapsid antibodies) were were not included in this study. All study participants provided written informed consent. See **Supplementary Table 1** for further cohort details. Serum samples were collected by centrifugation and stored at -80°C until analysis.

### Cloning of SARS-CoV-2 spike constructs

The Omicron (BA.1 sublineage of B.1.1.529) spike construct was based on amino acid substitutions observed in initial isolates (EPI_ISL_6640916, EPI_ISL_6640917, EPI_ISL_6640919) and carries the following changes in the spike protein compared to the Wu01 (EPI_ISL_406716) strain: A67V, Δ69-70, T95I, G142D, Δ143-145, N211I, Δ212, ins215EPE, G339D, S371L, S373P, S375F, K417N, N440K, G446S, S477N, E484A, Q493R, G496S, Q498R, N501Y, Y505H, T547K, D614G, H655Y, N679K, P681H N764K, D796Y, N856K, Q954H, N969K, and L981F. Codon-optimized overlapping gene fragments (Thermo Fisher) were assembled and cloned into the pCDNA3.1/V5-HisTOPO vector (Thermo Fisher) using the NEBuilder HiFi DNA Assembly Kit (New England Biolabs). The Alpha (B.1.1.7, with Δ69-70, Δ144, N501Y, A570D, D614G, P681H, T716I, S982A, and D1118H changes), Beta (B.1.351, with D80A, D215G, Δ242-4, K417N, E484K, N501Y, A570D, D614G, and A701V changes), and Delta (B.1.617.2, with T19R, G142D, Δ156-7, R158G, L452R, T478K, D614G, P681R, and D950N changes) spike variants were produced by PCR-induced mutagenesis and assembly of PCR products using the NEBuilder HiFi DNA Assembly Kit. All plasmid sequences were confirmed by sequencing.

### Pseudovirus neutralization assays

Pseudovirus neutralization assays were performed as previously described using a single-round infection lentivirus-based system^9,19^. Pseudovirus particles were generated in HEK293T cells by co-transfection of plasmids encoding for the SARS-CoV-2 spike protein, HIV-1 Tat, HIV-1 Gag/Pol, HIV-1 Rev, and luciferase followed by an IRES and ZsGreen using FuGENE 6 Transfection Reagent (Promega). Culture supernatants were changed to fresh medium 24 hours after transfection, and pseudovirus-containing supernatants were harvested at 48 h to 72 h post transfection. Following centrifugation and filtration (0.45 µm), pseudoviruses were stored at - 80°C until use. Pseudoviruses were titrated by infecting 293T-ACE2 cells and luciferase activity was determined following a 48-hour incubation period at 37°C and 5% CO_2_ by addition of luciferin/lysis buffer (10 mM MgCl_2_, 0.3 mM ATP, 0.5 mM Coenzyme A, 17 mM IGEPAL (all Sigma-Aldrich), and 1 mM D-Luciferin (GoldBio) in Tris-HCL) using a microplate reader (Berthold). Pseudovirus dilutions resulting in an at least 1000-fold difference in relative light units (RLUs) between infected and non-infected 293T-ACE2 cells were used for neutralization assays.

Serum samples were heat-inactivated at 56°C for 45 min prior to use. Serial dilutions of serum (1:3 dilution series with a starting dilution of 1:10) and monoclonal antibodies (1:5 dilution series with a starting concentration of 10 µg/ml) were co-incubated with pseudovirus supernatants for one hour at 37°C prior to addition of 293T-ACE2 cells. Following a 48-hour incubation at 37°C and 5% CO_2_, luciferase activity was determined as described above. Serum samples were tested in single dilution series and monoclonal antibodies were tested in duplicates. Background RLUs of non-infected cells were subtracted, and the serum ID_50_s and antibody IC_50_s were determined as the serum dilution and antibody concentration resulting in a 50% RLU reduction compared to virus-infected untreated controls cells using a non-linear fit model to plot an agonist vs. normalized dose response curve with variable slope using the least squares fitting method in GraphPad Prism 7.0 (GraphPad).

### Statistical methods

Comparison of neutralizing titers between different variants was performed using a two-tailed Wilcoxon matched-pairs signed rank test. Serum samples that did not achieve 50% inhibition at the lowest tested dilution of 10 (lower limit of quantification, LLOQ) were imputed to ½ of the LLOQ (ID_50_=5) for graphical representation and statistical evaluation.

### Data availability

All data are included in the figure, table, and supplement of this manuscript.

### Code availability

Not applicable.

## Data Availability

All data are included in the figure, table, and supplement of this manuscript.

## Acknowledgments

We are grateful to all study subjects for their participation in our studies. We thank M. Augustin, F. Dewald, L. Gieselmann, B. Kurt, P. Mayer, N. Riet, S. Salomon, M. Schlotz, R. Stumpf, and H. Wüstenberg, as well as the members of the EICOV/COVIM Study Group for sample acquisition and processing: Y. Ahlgrimm, B. Al-Rim, K. Behn, N. Bethke, H. Bias, D. Briesemeister, C. Conrad, V.M. Corman, C. Dang-Heine, S. Dieckmann, D. Frey, J.-A. Gabelich, J. Gerdes, U. Gläser, A. Hetey, L. Hasler, E.T. Helbig, A. Horn, C. Hülso, S. Jentzsch, C. von Kalle, L. Kegel, A. Krannich, W. Koch, P. Kopankiewicz, P. Kroneberg, H. Le, M. Lisy, L.J. Lippert, C. Lüttke, P. de Macedo Gomes, B. Maeß, J. Michel, F. Münn, A. Nitsche, A.-M. Ollech, C. Peiser, C. Pley, A. Pioch, A. Richter, M. Rönnefarth, C. Rubisch, A. Sanchez Rezza, L. Ruby, I. Schellenberger, V. Schenkel, S. Schmidt, J. Schlesinger, G. Schwanitz, T. Schwarz, A.-S. Sinnigen, A. Stege, S. Steinbrecher, P. Stubbemann, C. Thibeault, D. Treue, and S. Zvorc. This work was supported by grants from COVIM: „NaFoUniMedCovid19“ (FKZ: 01KX2021) (to L.E.S. and F.Kl.), the Federal Institute for Drugs and Medical Devices (V-2021.3 / 1503_68403 / 2021-2022) (to F.Ku. and L.E.S.), by the German Center for Infection Research (DZIF) (to F.Kl.), and the DFG CRC1279 and CRC1310 (to F.Kl.).

## Author contributions

Conceptualization, H.G., K.V., and F.Kl; Methodology, K.V. and F.Kl.; Investigation, H.G. and K.V.; Resources, P.T-L., D.H., P.S., C.L., F.Ku., and L.E.S.; Formal Analysis, H.G., K.V., and F.Kl. Writing - Original Draft, H.G., K.V., L.E.S., and F.Kl.; Writing - Review & Editing, all authors; Visualization, H.G., K.V., and F.Kl.; Supervision, F.Ku., L.E.S., and F.Kl.; Funding Acquisition, F.Ku., L.E.S. and F.Kl.

## Competing interests

H.G., K.V., and F.Kl. are listed as inventors on pending patent application(s) on SARS-CoV-2-neutralizing antibodies filed by the University of Cologne.

## Notes

### Author Declarations

COVID-19-convalescent samples were obtained under protocols approved by the ethics committee (EC) of the Medical Faculty of the University of Cologne (16-054 and 20-1187). Samples from non-infected vaccinated individuals were obtained under protocols (EICOV, COVIMMUNIZE, and COVIM) approved by the EC of Charite - Universitaetsmedizin Berlin (EA4/245/20 and EA4/244/20; EICOV and COVIMMUNIZE), and by the Federal Institute for Vaccines and Biomedicines (Paul Ehrlich Institute) and the EC of the state of Berlin (COVIM).

## References

1. Krammer, F. SARS-CoV-2 vaccines in development. Nature 586, 516–527 (2020).

2. Madhi, S.A., et al. Efficacy of the ChAdOx1 nCoV-19 Covid-19 Vaccine against the B.1.351 Variant. N Engl J Med 384, 1885–1898 (2021).

3. Planas, D., et al. Reduced sensitivity of SARS-CoV-2 variant Delta to antibody neutralization. Nature 596, 276–280 (2021).

4. UK Health Security Agency. SARS-CoV-2 variants of concern and variants under investigation in England. Technical briefing 31. (2021).

5. Pulliam, J.R.C., et al. Increased risk of SARS-CoV-2 reinfection associated with emergence of the Omicron variant in South Africa. medRxiv (2021).

6. Schmidt, F., et al. High genetic barrier to SARS-CoV-2 polyclonal neutralizing antibody escape. Nature (2021).

7. Greaney, A.J., et al. Comprehensive mapping of mutations in the SARS-CoV-2 receptor-binding domain that affect recognition by polyclonal human plasma antibodies. Cell Host Microbe 29, 463–476 e466 (2021).

8. Tober-Lau, P., et al. Long-term immunogenicity of BNT162b2 vaccination in older people and younger health-care workers. The Lancet Respiratory Medicine 9, e104–e105 (2021).

9. Vanshylla, K., et al. Kinetics and correlates of the neutralizing antibody response to SARS-CoV-2 infection in humans. Cell Host Microbe 29, 917–929 e914 (2021).

10. O’Brien, M.P., et al. Subcutaneous REGEN-COV Antibody Combination to Prevent Covid-19. N Engl J Med 385, 1184–1195 (2021).

11. Dougan, M., et al. Bamlanivimab plus Etesevimab in Mild or Moderate Covid-19. N Engl J Med 385, 1382–1392 (2021).

12. Gupta, A., et al. Early Treatment for Covid-19 with SARS-CoV-2 Neutralizing Antibody Sotrovimab. N Engl J Med 385, 1941–1950 (2021).

13. Khoury, D.S., et al. Neutralizing antibody levels are highly predictive of immune protection from symptomatic SARS-CoV-2 infection. Nat Med 27, 1205–1211 (2021).

14. Sette, A. & Crotty, S. Adaptive immunity to SARS-CoV-2 and COVID-19. Cell 184, 861–880 (2021).

15. Goel, R.R., et al. mRNA vaccines induce durable immune memory to SARS-CoV-2 and variants of concern. Science 374, abm0829 (2021).

16. Turner, J.S., et al. SARS-CoV-2 mRNA vaccines induce persistent human germinal centre responses. Nature 596, 109–113 (2021).

17. Muecksch, F., et al. Affinity maturation of SARS-CoV-2 neutralizing antibodies confers potency, breadth, and resilience to viral escape mutations. Immunity 54, 1853–1868 e1857 (2021).

18. Gaebler, C., et al. Evolution of antibody immunity to SARS-CoV-2. Nature 591, 639–644 (2021).

19. Crawford, K.H.D., et al. Protocol and Reagents for Pseudotyping Lentiviral Particles with SARS-CoV-2 Spike Protein for Neutralization Assays. Viruses 12(2020).

