## Supplementary Tables for "mRNA booster immunization elicits potent neutralizing serum activity against the SARS-CoV-2 Omicron variant"

**Supplementary Table 1.** *Study participant demographics.*

| **Vaccinated cohort** | |
|---|---|
| **Participants** - *n* | 30 |
| **Sex** | |
| Male - *n* (%) | 13 (43%) |
| Female - *n* (%) | 17 (57%) |
| **Age** - median years (range) | 49 (27-78) |
| **Vaccination received** | BNT162b2 |
| **Sampling time point -** median weeks (IQR; range) | |
| Early (after second dose) | 3.9 (3.7-4.2; 2.9-6.0) |
| Late (after second dose) | 21.0 (20.3-22.9; 18.7-31.1) |
| Booster (after second dose) | 40.6 (36.8-41.8; 29.7-44.0) |
| **Time between second and third dose -** median weeks (IQR; range) | 36.6 (32.2-38.5; 26.9-40.9) |

| **Convalescent cohort** | |
|---|---|
| **Participants** - *n* | 30 |
| **Sex** | |
| Male - *n* (%) | 14 (47%) |
| Female - *n* (%) | 16 (53%) |
| **Age** - median years (range) | 52 (22-68) |
| **Time period of SARS-CoV-2 infection** | February - March 2020 |
| **Disease severity** - *n* | |
| Mild symptoms | 29 |
| Hospitalized | 1 |
| **Sampling time point** - median weeks (IQR; range) | |
| Early (after disease onset) | 6.0 (4.7-7.0; 3.6-10) |
| Late (after disease onset) | 52.9 (51.4-56.8; 49.4-59.4) |
| Hybrid (after disease onset) | 66.8 (65.3-68.4; 62.4-71.4) |
| **Vaccination received** | BNT162b2 |
| **Time between infection and vaccination** - median weeks (IQR; range) | 60.9 (58.9-61.8; 52.4-66.1) |

**Supplementary Table 2.** *Serum neutralizing activity against Wu01 and Omicron.*

| Vaccinated cohort | | | | | | |
|---|---|---|---|---|---|---|
| | Wu01 Serum $ID_{50}$ | | | Omicron Serum $ID_{50}$ | | |
| Study ID | Early | Late | Boost | Early | Late | Boost |
| Pt. #001 | 41 | <10 | 5,079 | <10 | <10 | 324 |
| Pt. #002 | 37 | 14 | 2,537 | <10 | <10 | 624 |
| Pt. #003 | 293 | 28 | 2,545 | <10 | <10 | 516 |
| Pt. #004 | 242 | 75 | 1,849 | <10 | <10 | 2,410 |
| Pt. #005 | 264 | 41 | 7,706 | <10 | <10 | 2,907 |
| Pt. #006 | 1,749 | 339 | 11,807 | 12 | <10 | 2,872 |
| Pt. #007 | 689 | 231 | 6,149 | <10 | 11 | 1,485 |
| Pt. #008 | 588 | 267 | 9,734 | <10 | <10 | 1,690 |
| Pt. #009 | 254 | 115 | 2,948 | <10 | <10 | 756 |
| Pt. #010 | 1,478 | 467 | 22,912 | 28 | <10 | 881 |
| Pt. #011 | 1,342 | 372 | 22,833 | 33 | 37 | 5,510 |
| Pt. #012 | 726 | 94 | 6,211 | <10 | <10 | 1,148 |
| Pt. #013 | 963 | 430 | 6,909 | 35 | 44 | 1,741 |
| Pt. #014 | 3,336 | 817 | 11,718 | 114 | 21 | 3,050 |
| Pt. #015 | 1,006 | 165 | 5,567 | 15 | <10 | 428 |
| Pt. #016 | 521 | 101 | 10,154 | <10 | <10 | 2,484 |
| Pt. #017 | 823 | 54 | 3,077 | <10 | <10 | 1,185 |
| Pt. #018 | 1,128 | 265 | 8,731 | <10 | 11 | 924 |
| Pt. #019 | 1,008 | 239 | 6,461 | <10 | <10 | 601 |
| Pt. #020 | 1,334 | 225 | 6,269 | <10 | 14 | 450 |
| Pt. #021 | 695 | 535 | 11,128 | <10 | 36 | 2,645 |
| Pt. #022 | 1,600 | 326 | 19,819 | 49 | 134 | 7,851 |
| Pt. #023 | 294 | 227 | 7,643 | <10 | <10 | 868 |
| Pt. #024 | 311 | 98 | 11,463 | 23 | 53 | 3,514 |
| Pt. #025 | 474 | 160 | 3,768 | <10 | <10 | 1,312 |
| Pt. #026 | 551 | 327 | 6,078 | <10 | <10 | 1,571 |
| Pt. #027 | 831 | 112 | 6,317 | 10 | <10 | 625 |
| Pt. #028 | 408 | 118 | 7,307 | <10 | 23 | 1,454 |
| Pt. #029 | 270 | 93 | 3,676 | <10 | 21 | 2,239 |
| Pt. #030 | 352 | 68 | 609 | <10 | <10 | 21 |

| Convalescent cohort | | | | | | |
|---|---|---|---|---|---|---|
| | Wu01 Serum $ID_{50}$ | | | Omicron Serum $ID_{50}$ | | |
| Study ID | Early | Late | Hybrid | Early | Late | Hybrid |
| R014 | 330 | <10 | 5,178 | <10 | <10 | 141 |
| R047 | 2,927 | 566 | 8,932 | <10 | <10 | 1,657 |
| R056 | 1,355 | 88 | 4,137 | <10 | <10 | 74 |
| R082 | 3,144 | 108 | 7,948 | <10 | <10 | 2,188 |
| R090 | 623 | 119 | 8,650 | <10 | <10 | 4,528 |
| R102 | 862 | 90 | 27,730 | <10 | <10 | 3,187 |
| R137 | 204 | 141 | 8,581 | <10 | <10 | 430 |
| R164 | 669 | 70 | 5,818 | <10 | <10 | 725 |
| R212 | 217 | 80 | 5,347 | <10 | <10 | 343 |
| R238 | 731 | 140 | 14,687 | 87 | 76 | 11,001 |
| R244 | 2,505 | 119 | 24,552 | <10 | <10 | 5,843 |
| R247 | 1,032 | 142 | 13,058 | <10 | 14 | 2,034 |
| R256 | 8,735 | 397 | 23,738 | <10 | 31 | 16,570 |
| R257 | 394 | 353 | 9,332 | <10 | 20 | 907 |
| R285 | 264 | 75 | 3,450 | <10 | <10 | 3,888 |
| R289 | 217 | 153 | 7,964 | <10 | <10 | 3,888 |
| R297 | 547 | 89 | 30,227 | <10 | <10 | 3,397 |
| R299 | 73 | 30 | 5,709 | <10 | <10 | 1,233 |
| R317 | 438 | 133 | 9,163 | <10 | <10 | 2,205 |
| R399 | 76 | 26 | 2,338 | <10 | <10 | 474 |
| R401 | 59 | 38 | 4,610 | <10 | <10 | 1,278 |
| R434 | 473 | 137 | 5,264 | <10 | <10 | 430 |
| R440 | 1,250 | 381 | 21,847 | <10 | 11 | 1,375 |
| R458 | 171 | 14 | 2,553 | <10 | <10 | 874 |
| R484* | 971 | 289 | 15,642 | 17 | 34 | 8,297 |
| R491 | 421 | 99 | 7,680 | <10 | 17 | 806 |
| R527 | 37 | 18 | 2,979 | <10 | <10 | 400 |
| R715 | 91 | 55 | 6,999 | <10 | <10 | 2,381 |
| R749 | 11,008 | 95 | 4,579 | 79 | 31 | 8,028 |
| R849 | 219 | 160 | 7,640 | <10 | 11 | 4,546 |

| Serum $ID_{50}$ | >2,500 | 250-2,500 | 50-250 | 10-50 | <10 |
|---|---|---|---|---|---|

* Participant R484 received two BNT162b2 vaccinations three weeks apart.
